# Obesity’s Disparate Impact on COVID-19 Outcomes in Asian American Patients with Cancer

**DOI:** 10.1101/2025.05.14.25327577

**Authors:** Ryan H. Nguyen, Pankil K. Shah, Sanjay Mishra, Julie Fu, Sonya Reid, Jessica E. Hawley, Clara Hwang, Sunny R.K. Singh, Na Tosha Gatson, Narjust Florez, David Chism, Oscar K. Serrano, Babar Bashir, Gayathri Nagaraj, Rana R. Mckay, Brandon Hayes-Lattin, Vadim S Koshkin, Jared D. Acoba, Olga Zamulko, Christoper L. Geiger, Jeremy L. Warner, Dimpy P. Shah, Neeta K. Venepalli, the COVID-19 and Cancer Consortium

**Author notes:** Co-first authors (Ryan H. Nguyen, Pankil K. Shah). Co-last authors (Dimpy P. Shah, Neeta K. Venepalli). <u>Correspondence</u>: Ryan Nguyen, DO, University of Illinois at Chicago, 840 S Wood St, 820-E CSB, MC 713, Chicago, IL 60612; Pankil K. Shah, MD, PhD, UTHealth San Antonio, 7703 Floyd Curl Drive, San Antonio, TX, 78229, takes full responsibility for the manuscript as a whole. <u>Competing Interests</u> The authors declare no competing financial interests.

## Abstract

Data on COVID-19 outcomes in Asian Americans and Pacific Islanders (AAPI) are lacking. We analyzed data from 6,244 patients from the COVID-19 and Cancer Consortium, including 6.0% AAPI patients, to examine disparities in outcomes following acute COVID-19. Despite AAPI patients having lower median BMI than non-Hispanic White (NHW) patients and higher diabetes rates, both groups showed similar 30-day mortality and COVID-19 outcomes. In multivariable analyses, morbid obesity (BMI >35 kg/m^2^) in AAPI patients was associated with significantly higher 30-day mortality (OR 4.96; 95% CI 1.78 – 13.88) and COVID-19 severity (OR 2.6; 95% CI 1.26 – 5.35); underscoring the differential impact of obesity in AAPI patients.

## Introduction

Asian Americans and Pacific Islanders (AAPI) comprise one of the fastest-growing racial and ethnic groups in the US^1^. Yet, there is a paucity of published data on COVID-19-specific outcomes in this population.^2^ Immunocompromised patients, including those with cancer, are at higher risk of COVID-19 infection and severe disease.^3^ This novel study examines clinical characteristics and factors associated with disparate outcomes in AAPI patients compared to NHW patients with cancer and COVID-19.

## Methods

### Study Population and Data Sources

The COVID-19 and Cancer Consortium (CCC19) registry captures rapid, systematic, and detailed clinical characteristics, course of illness, and outcomes of COVID-19 among patients with cancer. In this registry-based, retrospective cohort study, we examined all reports of laboratory-confirmed SARS-CoV-2 infection submitted to the CCC19 registry between March 17, 2020, and December 31, 2021, for US residents with current or past diagnosis of cancer and Non-Hispanic AAPI or Non-Hispanic White (NHW) race and ethnicity.

We described demographics, comorbidities, tumor characteristics, and baseline severity of infection, stratified by AAPI and NHW groups. We also assessed outcomes such as 30-day mortality rate, mechanical ventilation use (MV), intensive care unit admission (ICU), and hospitalization and a 5-level ordinal scale of COVID-19 severity based on the patient’s most severe disease status: (1) none of the complications listed here, (2) hospital admission, (3) intensive care unit admission, (4), mechanical ventilation use, and (5) death from any cause. All clinical outcomes were all-cause in adult patients with cancer and laboratory-confirmed SARS-CoV-2. Using multiple imputations (m=10) to account for missingness of less than 5% in independent variables, we built unadjusted, minimally adjusted, and fully adjusted logistic regression models to estimate the association of the prior selected demographic and clinical variables on 30-day all-cause mortality and the ordinal severity outcome. The e-value-based sensitivity analysis was used to assess the potential effect of unmeasured confounding^4^. The analysis was conducted using R version 4.4.0 (2024). This study (NCT04354701) was approved by the Vanderbilt University Medical Center institutional review board and participating sites. A list of participating sites and the data dictionary are available at www.ccc19.org.

## Results

### Patient characteristics

From 6244 patients included in the analysis, 372 (6.0%) were AAPI, and 5872 (94.0%) were NHW (table 1). The median (IQR) age at COVID-19 diagnosis was 64 (53-73) for AAPI and 67 (57-77) for NHW. Female sex was 55.6% for AAPI and 51.1% for NHW. At the time of COVID-19 diagnosis, AAPI had lower rate of comorbidities, including pulmonary (14.5% vs 21.1%), cardiovascular (21.8% vs. 33.4%), and lower BMI measured as kg/m^2^ (median 25.1 vs 28.1) and smoking history (current or former smoker 30.4% vs. 49.0%) while having higher rates of diabetes (32.0% vs. 22.9%) compared with NHW. Tumor type, cancer status, type of cancer treatment, and timing of systemic therapy relative to COVID-19 diagnosis were similar between the two groups. AAPI were primarily located in the US West (44.6%) and Northeast (32.3%) regions in urban (42.5%) and suburban (34.9%) areas.

**Table 1.** Baseline characteristics of AAPI and NHW with cancer diagnosed with COVID-19. AAPI = Asian American and Pacific Islander, NHW = non-Hispanic White, BMI = body mass index, ECOG PS = eastern cooperative oncology group performance status, US = United States.

|  | **NHW**<br>N = 5872 | **AAPI**<br>N = 372 | **Total Cohort**<br>N = 6244 |
| --- | --- | --- | --- |
| **Sex** |  |  |  |
| Female | 2998 (51.1%) | 207 (55.6%) | 3205 (51.3%) |
| Male | 2872 (48.9%) | 164 (44.1%) | 3036 (48.6%) |
| Missing | 2 (0.0%) | 1 (0.3%) | 3 (0.0%) |
| **Age^** |  |  |  |
| Mean (SD) | 65.9 (14.5) | 62.0 (15.9) | 65.6 (14.6) |
| Median [Min, Max] | 67.0 [18.0, 90.0] | 64.0 [18.0, 90.0] | 67.0 [18.0, 90.0] |
| Median [Q1 - Q3] | 67.0 [57.0 - 77.0] | 64.0 [53.0 - 73.0] | 67.0 [57.0 - 76.0] |
| Missing | 3 (0.1%) | 0 (0%) | 3 (0.0%) |
| **Body Mass Index** |  |  |  |
| Mean (SD) | 29.1 (7.06) | 26.4 (6.62) | 29.0 (7.06) |
| Median [Min, Max] | 28.1 [11.9, 78.4] | 25.1 [14.5, 53.5] | 28.0 [11.9, 78.4] |
| Median [Q1 - Q3] | 28.1 [24.3 - 32.8] | 25.1 [22.1 - 29.5] | 28.0 [24.2 - 32.6] |
| Missing | 577 (9.8%) | 55 (14.8%) | 632 (10.1%) |
| **Modified Charlson Comorbidity Index** |  |  |  |
| Mean (SD) | 1.20 (1.64) | 1.09 (1.54) | 1.19 (1.64) |
| Median [Min, Max] | 1.00 [0, 10.0] | 0.500 [0, 8.00] | 1.00 [0, 10.0] |
| Median [Q1 - Q3] | 1.00 [0 - 2.00] | 0.500 [0 - 2.00] | 1.00 [0 - 2.00] |
| Missing | 49 (0.8%) | 2 (0.5%) | 51 (0.8%) |
| **Pulmonary Comorbidity** |  |  |  |
| No | 4608 (78.5%) | 316 (84.9%) | 4924 (78.9%) |
| Yes | 1240 (21.1%) | 54 (14.5%) | 1294 (20.7%) |
| Missing | 24 (0.4%) | 2 (0.5%) | 26 (0.4%) |
| **Diabetes** |  |  |  |
| No | 4479 (76.3%) | 251 (67.5%) | 4730 (75.8%) |
| Yes | 1344 (22.9%) | 119 (32.0%) | 1463 (23.4%) |
| Missing | 49 (0.8%) | 2 (0.5%) | 51 (0.8%) |
| **Cardiovascular Comorbidity** |  |  |  |
| No | 3860 (65.7%) | 289 (77.7%) | 4149 (66.4%) |
| Yes | 1963 (33.4%) | 81 (21.8%) | 2044 (32.7%) |
| Missing | 49 (0.8%) | 2 (0.5%) | 51 (0.8%) |
| **Renal Comorbidity** |  |  |  |
| No | 5039 (85.8%) | 325 (87.4%) | 5364 (85.9%) |
| Yes | 784 (13.4%) | 45 (12.1%) | 829 (13.3%) |
| Missing | 49 (0.9%) | 2 (0.5%) | 51 (0.8%) |
| **Smoking status** |  |  |  |
| Never | 2864 (48.8%) | 249 (66.9%) | 3113 (49.9%) |
| Current or Former | 2880 (49.0%) | 113 (30.4%) | 2993 (47.9%) |
| Unknown | 87 (1.5%) | 8 (2.2%) | 95 (1.5%) |
| Missing | 41 (0.7%) | 2 (0.5%) | 43 (0.7%) |
| **ECOG Performance status** |  |  |  |
| 0 | 1851 (31.5%) | 110 (29.6%) | 1961 (31.4%) |
| 1 | 1419 (24.2%) | 94 (25.3%) | 1513 (24.2%) |
| 2+ | 803 (13.7%) | 43 (11.6%) | 846 (13.5%) |
| Unknown | 1797 (30.6%) | 125 (33.6%) | 1922 (30.8%) |
| Missing | 2 (0.0%) | 0 (0%) | 2 (0.0%) |
| **Tumor Type** |  |  |  |
| Heme | 1006 (17.1%) | 58 (15.6%) | 1064 (17.0%) |
| Solid | 4059 (69.1%) | 279 (75.0%) | 4338 (69.5%) |
| Multiple | 807 (13.7%) | 35 (9.4%) | 842 (13.5%) |
| **Cancer Status** |  |  |  |
| Remission/NED | 2851 (48.6%) | 163 (43.8%) | 3014 (48.3%) |
| Active, responding or stable | 1619 (27.6%) | 89 (23.9%) | 1708 (27.4%) |
| Active, progressing | 737 (12.6%) | 49 (13.2%) | 786 (12.6%) |
| Unknown | 624 (10.6%) | 59 (15.9%) | 683 (10.9%) |
| Missing | 41 (0.7%) | 12 (3.2%) | 53 (0.8%) |
| **Timing of systemic therapy** |  |  |  |
| more than 3mth | 2730 (46.5%) | 178 (47.8%) | 2908 (46.6%) |
| 0-4 weeks | 1936 (33.0%) | 131 (35.2%) | 2067 (33.1%) |
| 1-3 months | 472 (8.0%) | 31 (8.3%) | 503 (8.1%) |
| Never or after COVID-19 | 493 (8.4%) | 24 (6.5%) | 517 (8.3%) |
| Missing | 242 (4.1%) | 8 (2.2%) | 249 (4.0%) |
| **Cytotoxic Cancer treatment** |  |  |  |
| No | 4719 (80.4%) | 291 (78.2%) | 5010 (80.2%) |
| Yes | 1058 (18.0%) | 77 (20.7%) | 1135 (18.2%) |
| Missing | 95 (1.6%) | 4 (1.1%) | 99 (1.6%) |
| **Targeted therapy** |  |  |  |
| No | 4913 (83.7%) | 307 (82.5%) | 5220 (83.6%) |
| Yes | 864 (14.7%) | 61 (16.4%) | 925 (14.8%) |
| Missing | 95 (1.6%) | 4 (1.1%) | 99 (1.6%) |
| **Endocrine therapy** |  |  |  |
| No | 5184 (88.3%) | 331 (89.0%) | 5515 (88.3%) |
| Yes | 593 (10.1%) | 37 (9.9%) | 630 (10.1%) |
| Missing | 95 (1.6%) | 4 (1.1%) | 99 (1.6%) |
| **Immunotherapy** |  |  |  |
| No | 5426 (92.4%) | 347 (93.3%) | 5773 (92.5%) |
| Yes | 351 (6.0%) | 21 (5.6%) | 372 (6.0%) |
| Missing | 95 (1.6%) | 4 (1.1%) | 99 (1.6%) |
| **Local therapy** |  |  |  |
| No | 5232 (89.1%) | 336 (90.3%) | 5568 (89.2%) |
| Yes | 545 (9.3%) | 32 (8.6%) | 577 (9.2%) |
| Missing | 95 (1.6%) | 4 (1.1%) | 99 (1.6%) |
| **Other cancer therapy** |  |  |  |
| No | 5668 (96.5%) | 362 (97.3%) | 6030 (96.6%) |
| Yes | 109 (1.9%) | 6 (1.6%) | 115 (1.8%) |
| Missing | 95 (1.6%) | 4 (1.1%) | 99 (1.6%) |
| **Region** |  |  |  |
| US Northeast | 2136 (36.4%) | 120 (32.3%) | 2256 (36.1%) |
| US Midwest | 1850 (31.5%) | 50 (13.4%) | 1900 (30.4%) |
| US South | 944 (16.1%) | 33 (8.9%) | 977 (15.6%) |
| US West | 886 (15.1%) | 166 (44.6%) | 1052 (16.8%) |
| Undesignated US | 56 (1.0%) | 3 (0.8%) | 59 (0.9%) |
| Non-US | 0 (0%) | 0 (0%) | 0 (0%) |
| **Residence** |  |  |  |
| Urban | 1614 (27.5%) | 158 (42.5%) | 1772 (28.4%) |
| Suburban | 2464 (42.0%) | 130 (34.9%) | 2594 (41.5%) |
| Rural | 714 (12.2%) | 9 (2.4%) | 723 (11.6%) |
| Other | 7 (0.1%) | 0 (0%) | 7 (0.1%) |
| Missing | 1073 (18.3%) | 75 (20.2%) | 1148 (18.4%) |
| **Insurance** |  |  |  |
| Medicare/Medicaid/Govt. | 2205 (37.6%) | 128 (34.4%) | 2333 (37.4%) |
| Private +/- other | 2096 (35.7%) | 95 (25.5%) | 2191 (35.1%) |
| Uninsured | 41 (0.7%) | 5 (1.3%) | 46 (0.7%) |
| Missing | 1530 (26.1%) | 144 (38.7%) | 1674 (26.8%) |
| **Treatment Center** |  |  |  |
| Academic Medical Center | 1370 (23.3%) | 61 (16.4%) | 1431 (22.9%) |
| Tertiary Care Center | 3046 (51.9%) | 280 (75.3%) | 3326 (53.3%) |
| Community Practice | 1455 (24.8%) | 31 (8.3%) | 1486 (23.8%) |
| Missing | 1 (0%) | 0 (0%) | 1 (0.0%) |
| **Year of COVID-19 diagnosis** |  |  |  |
| 2020 | 4670 (79.5%) | 322 (86.6%) | 4992 (79.9%) |
| 2021 | 1202 (20.5%) | 50 (13.4%) | 1252 (20.1%) |
| **Baseline COVID-19 Severity** |  |  |  |
| Mild | 3450 (58.8%) | 195 (52.4%) | 3645 (58.4%) |
| Moderate | 1923 (32.7%) | 146 (39.2%) | 2069 (33.1%) |
| Severe | 482 (8.2%) | 30 (8.1%) | 512 (8.2%) |
| Missing | 17 (0.3%) | 1 (0.3%) | 18 (0.3%) |

### COVID-19 Related Clinical Outcomes, Complications, Interventions

Clinical outcomes for 30-day mortality (11.0% vs 11.8%), mechanical ventilation [MV] (6.7% vs 8.4%), hospitalization (56.7% vs 53.2%), and ICU admission (14.0% vs 14.7%) were not statistically different between AAPI and NHW (supplementary figure 1). Consistent with the unadjusted and minimally adjusted models, the fully adjusted models showed that AAPI race and morbid obesity had a significant interaction effect that increased the odds of both 30-day mortality (OR, 4.96, 95% CI, 1.78-13.88) and COVID-19 severity (OR, 2.6, 95% CI, 1.26-5.35) outcomes (table 2). The E-value-based sensitivity analysis for the 30-day mortality outcome showed that an unadjusted factor would need to be associated with both mortality and being morbidly obese AAPI with an OR of at least 2.95 to attenuate the significance of the observed association. Similarly, the E-value-based sensitivity analysis for the ordinal severity outcome showed that an unadjusted factor would need to be associated with both severity and being morbidly obese AAPI with an OR of at least 1.84 to attenuate the significance of the observed association. Most documented associations in the CCC19 cohort have been smaller than the reported E-value. In the stratified models, morbid obesity was associated with significantly higher 30-day mortality among AAPI patients (OR, 7.25, 95% CI, 2.28 - 22.99), but it was not associated with higher mortality among NHW patients (OR, 0.92, 95% CI, 0.71 - 1.19) (Figure 1)^5^. Similarly, morbid obesity was also associated with significantly higher COVID-19 severity among AAPI patients (OR, 3.35, 95% CI, 1.55 - 7.2); however, not among NHW patients (OR, 0.92, 95% CI, 0.94 - 1.25). (Figure 1)

**Table 2:**
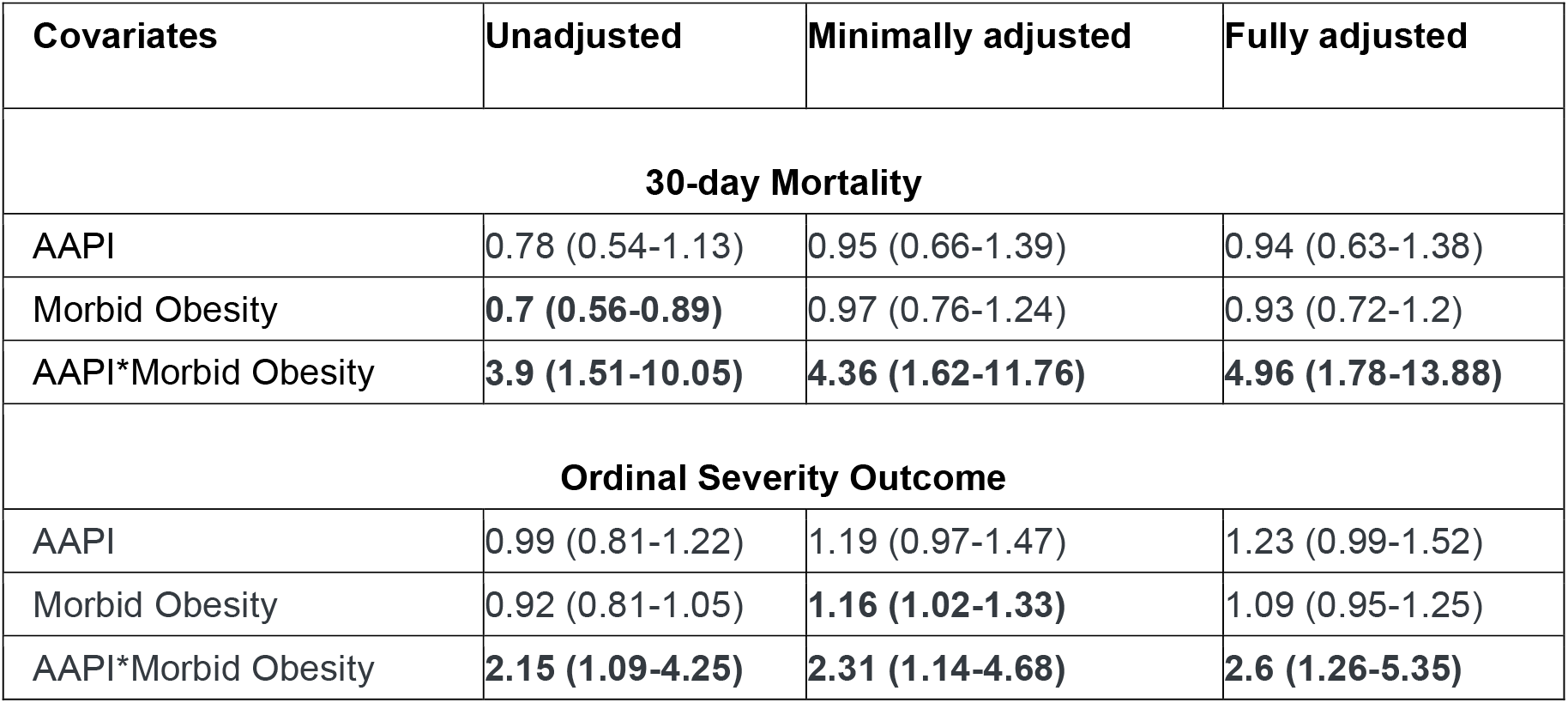
Multivariable regression models of AAPI and NHW with cancer diagnosed with COVID-19. AAPI = Asian American and Pacific Islander, NHW = Non-Hispanic White.

| Covariates | Unadjusted | Minimally adjusted | Fully adjusted |
| --- | --- | --- | --- |
| 30-day Mortality |  |  |  |
| AAPI | 0.78 (0.54-1.13) | 0.95 (0.66-1.39) | 0.94 (0.63-1.38) |
| Morbid Obesity | **0.7 (0.56-0.89)** | 0.97 (0.76-1.24) | 0.93 (0.72-1.2) |
| AAPI*Morbid Obesity | **3.9 (1.51-10.05)** | **4.36 (1.62-11.76)** | **4.96 (1.78-13.88)** |
| Ordinal Severity Outcome |  |  |  |
| AAPI | 0.99 (0.81-1.22) | 1.19 (0.97-1.47) | 1.23 (0.99-1.52) |
| Morbid Obesity | 0.92 (0.81-1.05) | **1.16 (1.02-1.33)** | 1.09 (0.95-1.25) |
| AAPI*Morbid Obesity | **2.15 (1.09-4.25)** | **2.31 (1.14-4.68)** | **2.6 (1.26-5.35)** |

**Figure 1:**
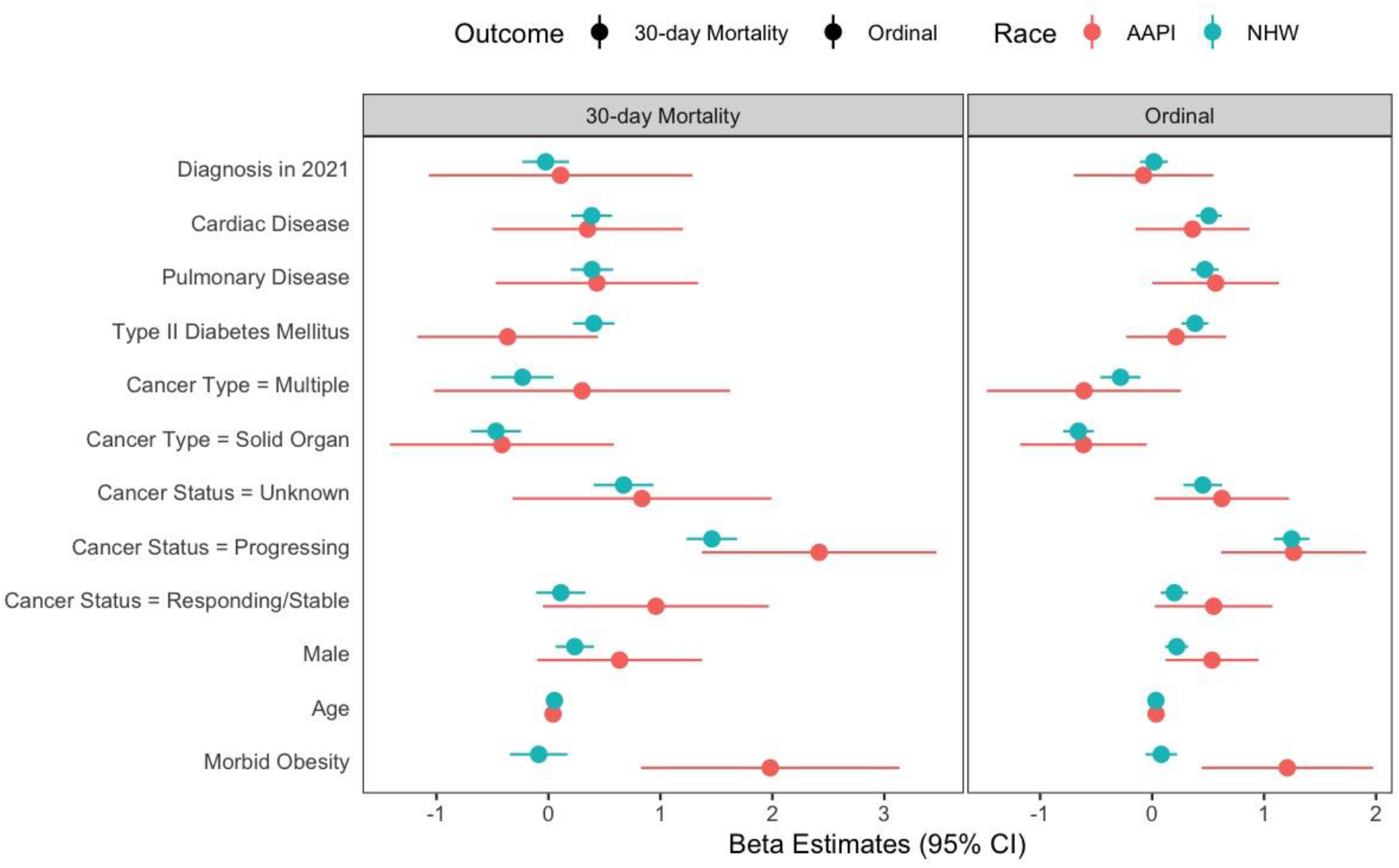
Forest plot of model coefficients for 30-day mortality and ordinal severity scale stratified by race

## Discussion

This retrospective cohort study is unique in evaluating COVID-19 illness and outcomes in the context of AAPI patients with cancer. Despite lower rates of comorbidities in AAPI patients, we found similar rates of COVID-19 clinical outcomes among the overall population of AAPI and NHW patients. However, morbid obesity had significant interaction with race and AAPI patients with morbid obesity experiencing significantly worse COVID-19 severity and 30-day mortality; while NHW patients with morbid obesity did not. The scarcity of AAPI-specific data highlights the need for further research to comprehend unique risks and vulnerabilities in this fast-growing demographic.

Cancer is the primary cause of death for both male and female AAPI individuals, while heart disease is the leading cause for all other major racial and ethnic groups in the US.^6,7^ Several factors contribute to cancer-related disparities among Asian Americans, including differences in infections, exposure to carcinogens, tumor biology^8^, and lower cancer screening rates^6^. This is despite Asian Americans having higher median household incomes, reduced poverty rates, and greater college education levels compared to the general US population.^1^ Data equity challenges also exist, stemming from the underreporting of AAPI individuals in health records due to inadequate collection and reporting of race and ethnicity. This issue is exacerbated by the aggregation of all Asian ethnic groups into a single race category or “other race.” ^9^

There is a dearth of AAPI-specific data related to COVID-19 outcomes. Early COVID-19 pandemic reports from San Francisco and New York showed higher than expected infection rates and higher mortality in AAPI compared to NHW individuals.^10,11^ National reports have suggested similar findings of higher mortality from COVID-19 among Asian Americans, but have been limited by lower proportions of Asian Americans in the databases compared to their relative U.S. population.^12^ The American Heart Association registry reported AAPI patients hospitalized with COVID-19 were significantly younger than NHW, due to higher rates of obesity and diabetes.^13^

Our study showed that AAPI patients with cancer were more likely to reside in urban settings and Western and Northeastern regions, be ‘never smokers’, and have fewer comorbidities except for diabetes. Although rates of obesity are lower in the AAPI population compared to other racial groups, the rates are rising, especially in 2nd and 3rd generations.^14^ Importantly, while we observed similar rates of 30-day mortality, hospitalizations, mechanical ventilation, and ICU admissions in AAPI and NHW patients, we noted morbid obesity in AAPI patients to significantly increase the risk of 30-day mortality and COVID-19 severity. These results suggest that morbid obesity may be associated with worse clinical consequences in AAPI patients.

AAPI patients have equivalent rates of type 2 diabetes and metabolic syndrome at lower BMI cutoffs compared to NHW patients, suggesting that the burden of obesity-related disease in AAPI populations may be underreported.^15^ Several organizations including the WHO recommend lower BMI cutoffs for obesity in Asian populations (BMI >25 or >27.5) to account for this increased risk.^16^ We analyzed clinical outcomes for NHW and AAPI patients by varying BMI cut-offs (25, 30, 35) and found the subgroup of AAPI patients with BMI >35 had the highest rates of 30-day mortality, hospitalization, ICU admission, and mechanical ventilation.

Etiologies for the increased amount of obesity related complications such as diabetes among AAPI patients include decreased physical activity, high carbohydrate intake, acculturation with higher rates of obesity and diabetes among successive generations in the US.^16^ Within AAPI grouping, some studies have reported heterogeneity in diabetes-related mortality, with Filipino patients experiencing the greatest burden.^17^ Additionally, when adjusted for age, BMI, and total fat mass, Chinese and South Asian patients have significantly more visceral adipose tissue than White counterparts.^18^ Additionally, obesity is recognized as a risk factor for hospitalization and death due to COVID-19 in the general population.^19^ While the exact reasons for the differential impact of obesity in AAPI compared to NHW patients remain unclear, our results highlight the importance of considering race- and ethnicity-specific factors when addressing health disparities.

This study is, to our knowledge, the largest cohort of AAPI patients with COVID-19 and cancer. Other strengths include representation from academic and community cancer centers across the US, and systematically collected granular data on characteristics, presentations, and outcomes of patients with cancer and COVID-19. Limitations include incomplete documentation of race and ethnicity in the electronic health records from which these data are derived, including a lack of subcategorization of race and ethnicity beyond Office of Management and Budget minimums. Cases for CCC19 were entered by self-selecting cancer centers and may not represent the broader population, however the distribution of sites throughout the country makes this the largest report of AAPI patients with cancer and COVID-19. Lastly, CCC19 does not collect detailed data on social determinants of health given the lack of these data in electronic health records. Further research is necessary on the specific cultural and social determinants of health factors which account for the differences in AAPI subgroups.

## Conclusions

Despite similar clinical outcomes related to COVID-19, morbid obesity was associated with substantially worse outcomes for AAPI patients compared to NHW patients. Additional investigation is required to uncover unique mechanisms driving the disparate impact of obesity in AAPI communities and to bridge the gaps in health data collection and reporting for this critically understudied population.

## Data Availability

Data from the CCC19 is available in aggregate, de-identified format at https://ccc19.org/data.html

## Acknowledgement

We thank all members of the CCC19 steering committee: Toni K Choueiri, MD; Dimitrios Farmakiotis, MD, FACP, FIDSA; Narjust Florez, MD; Corrie A. Painter, PhD; Solange Peters, MD, PhD; Petros Grivas, MD, PhD; Gilberto de Lima Lopes Jr, MD, MBA, FAMS, FASCO; Sonya A. Reid, MD, MPH; Dimpy P. Shah, MD, PhD; Michael Thompson, MD, PhD, FASCO; Jeremy L. Warner, MD, MS, FAMIA, FASCO; for their invaluable guidance of the CCC19 consortium.

